# Growing inequities by immigration group among older adults: Population-based analysis of access to primary care and return to in-person visits during the COVID-19 pandemic in British Columbia, Canada

**DOI:** 10.1101/2023.06.23.23291828

**Authors:** Cecilia Sierra-Heredia, Elmira Tayyar, Yasmin Bozorgi, Padmini Thakore, Selamawit Hagos, Ruth Carrillo, Stefanie Machado, Sandra Peterson, Shira Goldenberg, Mei-ling Wiedmeyer, Ruth Lavergne

**Affiliations:** Faculty of Health Sciences, Simon Fraser University, Burnaby, Canada; Centre for Gender & Sexual Health Equity, Vancouver, British Columbia, Canada; Centre for Health Services and Policy Research, University of British Columbia, British Columbia, Canada; School of Public Health, San Diego State University, San Diego, California, USA; Department of Family Practice, University of British Columbia, Vancouver, Canada; Department of Family Medicine, Faculty of Medicine, Dalhousie University, Nova Scotia, Canada

**Keywords:** Primary healthcare, immigration, language, access to primary care, virtual care, telemedicine

## Abstract

**Background:** The onset of the COVID-19 pandemic drove a rapid and widespread shift to virtual care, followed by a gradual return to in-person visits. Virtual visits may offer more convenient access to care for some, but others may experience challenges accessing care virtually, and some medical needs must be met in-person. Experiences of the shift to virtual care and benefits of in-person care may vary by immigration experience (immigration status and duration), official language level, and age. We examined use of virtual care and return to in-person visits in the Canadian province of British Columbia (BC), comparing patterns by age and across immigration groups, including length of time in Canada and language level (English) at time of arrival.

**Methods:** We used linked administrative health and immigration data to examine total primary care visits (virtual or in-person) and return to in-person visits during the COVID-19 pandemic (2019/20-2021/2) in BC. We examined the proportion of people with any primary care visits and with any in-person visit within each year as measures of access to primary care. We estimated the odds of any primary care visit and any in-person visit by immigration group and official language level assessed prior to arrival: non-immigrants, long-term immigrants, recent immigrants (<5 years) with high assessed English level and recent immigrants (<5 years) with low assessed English level, stratified by age.

**Results:** In general, changes in access to primary care (odds of any visit and odds of any in-person visit) were similar across immigration groups over the study period. However, we observed substantial disparities in access to primary care by immigration group among people aged 60+, particularly in recent immigrants with low official language level (0.42, 0.40-0.45). These disparities grew wider over the course of the pandemic.

**Conclusion:** Though among younger adults changes in access to primary care between 2019-2021 were similar across immigration groups, we observed significant and growing inequities among older adults, with particularly limited access among adults who immigrated recently and with low assessed English level. Targeted interventions to ensure acceptable, accessible care for older immigrants are needed.

## Background

Virtual care is defined as the provision of medical care, medical resources, or medical education, delivered remotely through the use of electronic information and technology (including phone, email, or videoconference communication) for the diagnosis, treatment and prevention of disease and injuries (1–3). Use of virtual care increased rapidly following the onset of the COVID-19 pandemic in March 2020. Data from the Canadian province of Ontario show widespread use of virtual care across the entire population, including vulnerable groups like older adults and lower-income patients (1). In Ontario, virtual care constituted 71.1% of all visits from January 1st to July 28, 2020, with higher proportions of virtual care visits among adults aged 65–74 years (73.4%) and those with the highest expected health care needs (73.1%) (4).

Though it appears use of virtual care is widespread (5,6) and initial data do not point to gaps in access for vulnerable groups (4), virtual platforms may not meet all patients’ needs. In addition, less is known about the return to in-person care for patients requiring it, following initial changes to health service delivery in the context of the COVID-19 pandemic. People who require assistance navigating care, require translation services, or have lower experience with or access to technology may face barriers in using virtual platforms (7,8) and in-person access may be particularly helpful for these populations. Whether they have been able to access in-person care since the COVID-19 pandemic was declared is unknown. In Manitoba, a study observed disproportionate reductions in care for children and adolescents from immigrant and refugee families and low socioeconomic status (9) compared to Canadian-born children and adolescents, but less is known about impacts across the adult population.

In Canada, immigrants face barriers accessing health care in general (10), and particularly if they have more recently arrived in Canada (11). Immigrants may also face challenges navigating access to virtual care, and obtaining care if one of the two official languages in Canada (English or French) is not their preference for health care communication (12,13). Integration of translation in virtual platforms can be and has been challenging (14) and in the presence of discordant linguistic preferences there is an increased risk of diagnosis delays or errors, delayed care, and inappropriate treatments (15). Factors like suboptimal access to internet networks or software (7), and lower levels of ehealth literacy (the individual’s and community’s ability to access and apply information about health with digital services, (8) have also been shown to be significant barriers to accessing virtual health care (1).

In the context of the COVID-19 pandemic, it may be that people for whom virtual care was not optimal had no other options, and there is limited information worldwide about the return to in-person care when pandemic restrictions eased (16,17). We thus compare any primary care use and access to any in-person care by immigration groups and official language level, stratifying by age, over 2019/20 to 2021/2, with a focus on return to in-person care following the initial wave of COVID-19, once safety protocols were in place to support this return in later 2020.

## Methods

### Study design

This longitudinal, population-based study used linked administrative health and immigration data to examine total primary care visits (virtual or in-person) and return to in-person primary care visits in British Columbia (BC), Canada between the years of 2019/20 to 2021/2.

### Setting

Primary care is publicly funded for people who qualify for provincial medical insurance in British Columbia (BC). This excludes people who have some forms of temporary status or who do not have current legal immigration status. In BC, the federal ministry of Immigration, Refugees and Citizenship Canada (IRCC) controls immigration and movement across federal borders by issuing travel documents and screening potential permanent and temporary residents (18); the federal and provincial borders of BC were defined on the lands of more than 200 Indigenous nations through historical and ongoing colonial processes (19).

Primary care in BC is paid predominantly via fee-for-service payments to physicians, or alternate payment plans for physicians and nurse practitioners who shadow bill (or do encounter coding) for visits. Though the ability to bill for primary care delivered virtually has existed in British Columbia since 2013, virtual visits remained only a small portion of total primary care prior to the COVID-19 pandemic (20,21), as in many other settings globally (2,6). After the COVID-19 pandemic was declared on March 11, 2020 (22) and public health measures were put in place to reduce the risk of transmission (23), virtual care was identified as an alternative to in-person care that could address patient needs, while also reducing risk of transmission (24,25). Adoption of virtual care subsequently increased rapidly in BC and nationally (1,6). On March 16, 2020, the BC Ministry of Health announced enhanced availability of physician compensation for virtual care services (26). On May 21, 2020 following guidelines from the Provincial Health Officer, the College of Physicians and Surgeons of BC (CPSBC), WorkSafeBC, and BC Centres for Disease Control, the provincial physician association, Doctors of BC, published recommendations for expanding safe in-person care (27). While no timelines were set for expanding safe in-person care, the recommendations included guidance on clinics developing a COVID safety plan, such as adequate supply of personal protective equipment, safety measures for patients, and changes to clinic hours (27). However, between April and September 2020, 86% of patients in BC were still accessing primary care virtually (28). Fifteen months later, as BC’s vaccination rates reached 80%, on September 3, 2021 provincial health officials (the Assistant Deputy Minister of the Primary Care Division, the Provincial Health Officer, and the Registrar and CEO CPSBC) released a letter urging physicians to return to in-person care (29).

### Data

We accessed linked, population-based, administrative data through Population Data BC, including the Medical Services Plan (MSP) registry file/Central Demographics File (30), physician payments (31), and hospitalizations (used to derive measures of comorbidity) (32). Data used also included Immigration, Refugees and Citizenship Canada (IRCC)-PR data (33) and the MSP Residency Status data (30). Access to data provided by the Data Steward(s) is subject to approval, but can be requested for research projects through the Data Steward(s) or their designated service providers. All inferences, opinions, and conclusions drawn in this publication are those of the author(s), and do not reflect the opinions or policies of the Data Steward(s).

### Population

The study population included all people aged 20 (as of April 1, 2019) and older registered for British Columbia Medical Services Plan (MSP) for at least 75% of each year of 2019/20, 2020/1, 2021/2. We excluded people identified in the MSP registry as visitors, diplomats, and on working holiday visas in BC as their usual place of residence is outside of Canada. People identified in the MSP registry as having temporary status who do not appear in (IRCC) data were excluded because key information about language and length of time in Canada was collected only for permanent residents during their application process. People who died or who were in long-term care were excluded.

### Measures

Our primary outcomes of interest were any visit with a family physician or nurse practitioner (defined as a unique combination of patient, provider and date, regardless of fee items billed, for visits that occurred in the community) and any in-person primary care visit (the subset of visits that did not include fee items specific to virtual care). We chose to focus on having any visit within the year rather than visit volume as a marker of any contact with primary care. This is particularly important in the context of the pandemic, given the potential for people to have lost contact with healthcare, and who may lack a regular place of care or first point of access (34,35).

We included four immigration groups based on records from IRCC: *Non-immigrant/long-term residents* of Canada include people with no record in IRCC data going back to 1985, the starting date of the dataset we used. *Long-term immigrants* include people in IRCC data who were in Canada 5 years or more as of April 1, 2019. *Recent immigrants* include people in IRCC data who were in Canada less than 5 years as of April 1, 2019. This group was further divided into *people with high English level* (assessed prior to arrival), and *people with low English level*. As official language level is documented by IRCC prior to arrival, we chose to disaggregate by language level only among people in Canada less than 5 years. This reduced potential misclassification as language level changes over time, while allowing us to explore the combined effect of non-English language preferences and lack of familiarity with healthcare in BC.

The MSP registration form contains a variable labeled “Gender” with the options “M” and “F” provided (presumed to be abbreviations of the sexes “male” and “female”). Whether responses reflect gender, sex assigned at birth or legal sex cannot be determined. We refer to this variable as “administrative sex”. Neighbourhood income quintiles were determined based on census enumeration area of residence, assigned using the Postal Code Conversion File (PCCF+) (36). We used the Statistics Canada Statistical Area Classification Metropolitan Influences Zones to group metropolitan areas (census metropolitan areas), small urban areas (census agglomerations) and rural/remote settings (areas with strong to no metropolitan influence) (37). Regional health authority of residence and the number of comorbidities were measured using rolling two year periods for the Charlson index (38).

### Analysis

We described the study population by age and immigration group and plotted the percentage of the population with any primary care visits and in-person visits over the study period (6 month intervals), stratified by age and immigration group. We then explored factors (i.e. immigration status, age, administrative sex, rurality, income, and comorbidities) shaping whether or not people had any primary care visits or any in-person visits within each year. We used generalized linear models with binomial distribution and logit link to model odds of any primary care and any virtual care within each year. We reported unadjusted and adjusted odds of any primary care and any in-person primary care during pandemic years, relative to 2019, stratified by age. We included an interaction term between year and immigration group to test if any changes over the study period differ by immigration group. Adjusted models included 5-year age group, administrative sex, urban/rural residence, income quintile, and Charlson comorbidities entered as binary variables for each condition, as a measure of need for health care.

## Results

Table 1 shows the demographic characteristics of participants in the sample by age and immigration group. In all age groups, a higher percentage of recent immigrants with low English language level were “female” (58.0% for 20-39, 58.5% for 40-59, and 58.9 % for 60+). In all age groups, higher percentages of both recent and long-term immigrants lived in metropolitan settings (86.6-96.1%), than non-immigrants/long-term residents (immigrants who arrived before 1985) (58.2-64.2%). In all age groups, a disproportionate percentage of immigrants lived in the lowest income quintiles, though which immigration groups were most concentrated in the lowest income neighbourhoods varies by age. Among people aged 20-39 and 40-59, higher percentages of recent immigrants with low English language level lived in the lowest income quintiles 34.2% and 29.4% respectively). Among people aged 60+ a higher percentage of long-term immigrants lived in the lowest income quintile (25.8%) than other immigration groups in this age group. The number of treated comorbidities increased with age, though patterns vary among immigration groups. Among people aged 20-39, recent immigrants with low English language level had the highest mean number of comorbidities. Among people aged 60+, non-immigrants had the highest mean number of comorbidities.

**Table 1a.** Study population characteristics by age and immigration group, British Columbia, Canada, 2019-2021 (n= 3,707,248)

|  | Age 20-39 |  |  |  | Age 40-59 |  |  |  | Age 60+ |  |  |  |
| --- | --- | --- | --- | --- | --- | --- | --- | --- | --- | --- | --- | --- |
| Characteristic | Non-immigrant<br>(or immigrant<br><1985) | Long-term<br>immigrant | Recent<br>immigrant,<br>high English<br>level | Recent<br>immigrant, low<br>official<br>language<br>proficiency | Non-immigrant<br>(or immigrant<br><1985) | Long-term<br>immigrant | Recent<br>immigrant,<br>high English<br>level | Recent<br>immigrant, low<br>official<br>language<br>proficiency | Non-immigrant<br>(or immigrant<br><1985) | Long-term<br>immigrant | Recent<br>immigrant,<br>high English<br>level | Recent<br>immigrant, low<br>official<br>language<br>proficiency |
| **N (%)** | 922,297<br>(24.9%) | 205,771<br>(5.6%) | 92,994<br>(2.5%) | 8,205<br>(0.2%) | 975,886<br>(26.3%) | 299,847<br>(8.1%) | 32,303<br>(0.9%) | 7,342<br>(0.2%) | 998,070<br>(26.9%) | 151,347<br>(4.1%) | 5,462<br>(0.1%) | 7,724<br>(0.2%) |
| **Age,\* mean**<br>**(SD)** | 29.6 (5.7) | 31.2 (5.6) | 31.2 (4.6) | 30.2 (5.5) | 50.2 (5.8) | 49.3 (5.6) | 46.4 (5.1) | 49.3 (5.8) | 70.3 (8.0) | 69.6 (8.3) | 67.9 (6.3) | 68.6 (6.6) |
| **Administrative sex, N (%) \*\*** |  |  |  |  |  |  |  |  |  |  |  |  |
| F | 461,008 (50.0) | 99,247 (48.2) | 50,424 (54.2) | 4,757 (58.0) | 500,905 (51.3) | 150,179 (50.1) | 16,995 (52.6) | 4,298 (58.5) | 536,880 (53.8) | 69,234 (45.7) | 2,726 (49.9) | 4,552 (58.9) |
| M | 461,286 (50.0) | 106,524 (51.8) | 42,570 (45.8) | 3,448 (42.0) | 474,971 (48.7) | 149,668 (49.9) | 15,308 (47.4) | 3,044 (41.5) | 461,114 (46.2) | 82,112 (54.3) | 2,736 (50.1) | 3,172 (41.1) |
| **Rurality, N (%)\*** |  |  |  |  |  |  |  |  |  |  |  |  |
| Metropolitan | 591,827 (64.2) | 190,170 (92.4) | 82,188 (88.4) | 7,645 (93.2) | 592,629 (60.7) | 274,148 (91.4) | 28,029 (86.8) | 6,986 (95.2) | 580,788 (58.2) | 139,743 (92.3) | 4,729 (86.6) | 7,424 (96.1) |
| Small urban | 209,232 (22.7) | 9,737 (4.7) | 6,385 (6.9) | 412 (5.0) | 228,883 (23.5) | 15,439 (5.1) | 2,715 (8.4) | 258 (3.5) | 247,536 (24.8) | 6,407 (4.2) | 412 (7.5) | 194 (2.5) |
| Rural/remote | 108,532 (11.8) | 4,001 (1.9) | 2,963 (3.2) | 105 (1.3) | 139,351 (14.3) | 8,104 (2.7) | 1,258 (3.9) | 69 (0.9) | 161,535 (16.2) | 4,481 (3.0) | 278 (5.1) | 82 (1.1) |
| Missing | 12,706 (1.4) | 1,863 (0.9) | 1,458 (1.6) | 43 (0.5) | 15,023 (1.5) | 2,156 (0.7) | 301 (0.9) | 29 (0.4) | 8,211 (0.8) | 716 (0.5) | 43 (0.8) | 24 (0.3) |
| **Neighbourhood income quintile (after tax), N (%)\*** |  |  |  |  |  |  |  |  |  |  |  |  |
| 1 (lowest) | 180,938 (19.6) | 49,258 (23.9) | 26,563 (28.6) | 2,809 (34.2) | 167,695 (17.2) | 66,182 (22.1) | 8,331 (25.8) | 2,156 (29.4) | 196,082 (19.6) | 39,092 (25.8) | 1,155 (21.1) | 1,827 (23.7) |
| 2 | 177,797 (19.3) | 48,656 (23.6) | 21,847 (23.5) | 2,231 (27.2) | 170,132 (17.4) | 68,742 (22.9) | 7,071 (21.9) | 1,785 (24.3) | 191,386 (19.2) | 37,683 (24.9) | 1,183 (21.7) | 1,936 (25.1) |
| 3 | 188,191 (20.4) | 42,742 (20.8) | 18,179 (19.5) | 1,557 (19.0) | 192,469 (19.7) | 61,494 (20.5) | 5,901 (18.3) | 1,342 (18.3) | 196,139 (19.7) | 30,634 (20.2) | 1,155 (21.1) | 1,537 (19.9) |
| 4 | 190,234 (20.6) | 35,628 (17.3) | 14,491 (15.6) | 940 (11.5) | 208,546 (21.4) | 52,946 (17.7) | 5,414 (16.8) | 978 (13.3) | 191,707 (19.2) | 23,341 (15.4) | 1,009 (18.5) | 1,303 (16.9) |
| 5 (highest) | 172,257 (18.7) | 27,622 (13.4) | 10,454 (11.2) | 625 (7.6) | 221,803 (22.7) | 48,305 (16.1) | 5,283 (16.4) | 1,052 (14.3) | 214,240 (21.5) | 19,866 (13.1) | 917 (16.8) | 1,097 (14.2) |
| Missing | 12,880 (1.4) | 1,865 (0.9) | 1,460 (1.6) | 43 (0.5) | 15,241 (1.6) | 2,178 (0.7) | 303 (0.9) | 29 (0.4) | 8,516 (0.9) | 731 (0.5) | 43 (0.8) | 24 (0.3) |
| **Comorbidities, N (%)\*** |  |  |  |  |  |  |  |  |  |  |  |  |
| 0 conditions | 795,098 (86.2) | 175,152 (85.1) | 80,542 (86.6) | 6,806 (82.9) | 714,260 (73.2) | 215,861 (72.0) | 24,981 (77.3) | 5,298 (72.2) | 491,150 (49.2) | 78,262 (51.7) | 3,031 (55.5) | 4,372 (56.6) |
| 1+ condition | 127,199 (13.8) | 30,619 (14.9) | 12,452 (13.4) | 1,399 (17.1) | 261,626 (26.8) | 83,986 (28.0) | 7,322 (22.7) | 2,044 (27.8) | 506,920 (50.8) | 73,085 (48.3) | 2,431 (44.5) | 3,352 (43.4) |
| # conditions,<br>Mean (SD) | 0.16 (0.42) | 0.17 (0.44) | 0.15 (0.40) | 0.20 (0.46) | 0.35 (0.66) | 0.35 (0.64) | 0.27 (0.56) | 0.35 (0.64) | 0.85 (1.08) | 0.76 (0.99) | 0.66 (0.91) | 0.63 (0.88) |

\*Values within 2019/2020 fiscal year.
\*\* Administrative sex was missing (or coded as "U" or "I") for 90 individuals in total.

**Table 1b.** Study population characteristics by age and immigration group, British Columbia, Canada, 2019-2021, Immigration class (people in IRCC data only), British Columbia, Canada, 2019-2021

| Characteristic. | Age 20-39 |  |  | Age 40-59 |  |  | Age 60+ |  |  |
| --- | --- | --- | --- | --- | --- | --- | --- | --- | --- |
|  | Long-term immigrant<br>n (%) | Recent immigrant,<br>high English level<br>n (%) | Recent immigrant,<br>low English level<br>n (%) | Long-term immigrant<br>n (%) | Recent immigrant,<br>high English level<br>n (%) | Recent immigrant,<br>low English level<br>n (%) | Long-term immigrant<br>n (%) | Recent immigrant,<br>high English level<br>n (%) | Recent immigrant,<br>low English level<br>n (%) |
| Economic, worker(caregiver) | 8,011 (3.9) | 8,251 (8.9) | 120 (1.5) | 12,572 (4.2) | 7,100 (22.0) | 123 (1.7) | 1,867-1,870<br>*(1.23-1.24) | 276 (5.1) | 12 (0.2) |
| Economic - provincial nominee & worker (other) | 87,911 (42.7) | 54,470 (58.6) | 856 (10.4) | 131,062 (43.7) | 15,323 (47.4) | 1,303 (17.7) | 43,672 (28.9) | 484 (8.9) | 54 (0.7) |
| Protected person | 6,995 (3.4) | 1,583 (1.7) | 423 (5.2) | 12,447 (4.2) | 815 (2.5) | 392 (5.3) | 7,292 (4.8) | 169 (3.1) | 161 (2.1) |
| Refugee, Blended sponsorship | 6 (0.0) | 94 (0.1) | 257 (3.1) | 8 (0.0) | 39 (0.1) | 103 (1.4) | ≤5 | ≤5 | 6 (0.1) |
| Refugee, Government assisted | 7,860 (3.8) | 368 (0.4) | 1,687 (20.6) | 9,836 (3.3) | 153 (0.5) | 929 (12.7) | 4,415 (2.9) | 20-23 (0.37-0.42)* | 153 (2.0) |
| Refugee, privately sponsored | 3,246 (1.6) | 1,291 (1.4) | 560 (6.8) | 4,814 (1.6) | 422 (1.3) | 397 (5.4) | 2,218 (1.5) | 62 (1.1) | 147 (1.9) |
| Sponsored family | 61,797 (30.0) | 25,869 (27.8) | 3,877 (47.3) | 99,272 (33.1) | 7,187 (22.2) | 2,627 (35.8) | 66,346 (43.8) | 4,265 (78.1) | 7,112 (92.1) |

\*Range presented so that surpassed values cannot be determined

The composition of immigration groups with respect to immigration class varied by age group. Among people aged 20-59, higher percentages of long-term and recent immigrants with high English level were in an economic immigration class, while people with low English language level include more refugees and sponsored family members. Among people aged 60+ with low English language level, over 92% were sponsored family members.

Across all age and immigration groups, the percentage of people with any primary care visit (in-person or virtual) fell in 2020, but then increased, though not to pre-pandemic levels (Figure 1). The percentage of people with in-person primary care visits fell dramatically in 2020, and then increased gradually, though not to pre-pandemic levels for any group. However, precise patterns differed by age and immigration group. Among people aged 20-39, a slightly higher percentage of people who immigrated recently with low English level have primary care visits, and this persists over the pandemic period. Among people aged 40-59 the percentage of people with primary care visits was similar by immigration group, though patterns change slightly over the course of the pandemic. Among people aged 60 and older, there were notable differences in service use by immigration groups. Within this group, non-immigrants had a higher percentage of visits, followed by long-term immigrants in Canada 5 or more years, recent immigrants in Canada (<5 years) with high English level, and finally recent immigrants in Canada with low official language proficiency. Differences between these groups appeared to grow wider over the course of the pandemic.

**Figure 1.**
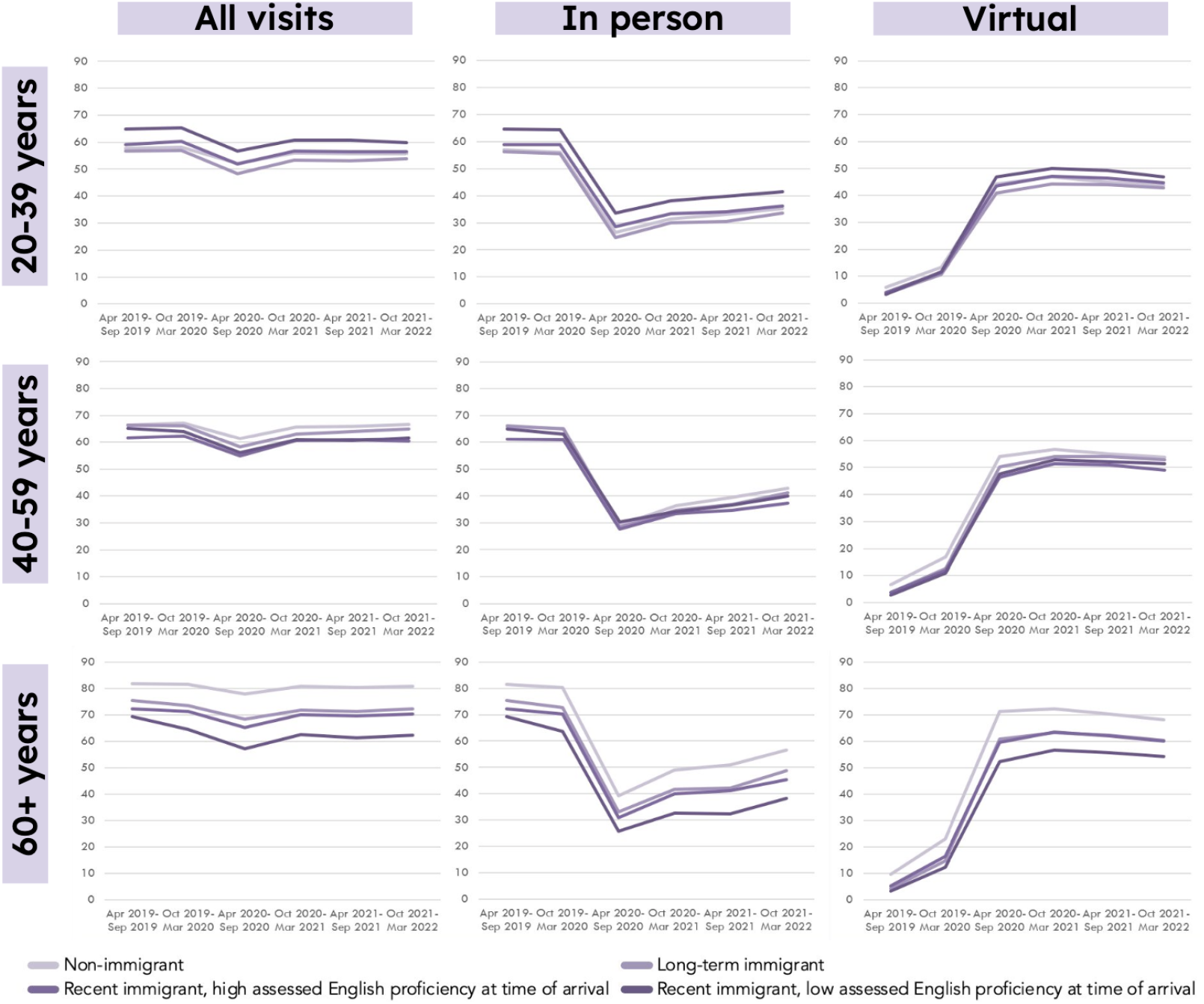
Percentage of the population with primary care visits (all visits and in-person only, over 6 month period) 2019/20-2021/2, British Columbia, Canada.

Adjusted Odds Ratio (AOR) and 95% Confidence Intervals (CI) of access to any primary care visits presented in Table 2 reinforce that among immigration groups, inequity in access to care centered around age. While recent immigrants with high English level had somewhat higher adjusted odds of any primary care visit among people aged 20-39 (1.05, 1.03-1.06) compared to same-age non-immigrants, among people aged 40-59, recent immigrants with high English level had lower adjusted odds of any primary care visit (0.87, 0.84-0.89), and odds were much lower among people aged 60+ (0.58, 0.53-0.62). Among recent immigrants with low English language level, adjusted odds ratios of access to any primary care visits were more extreme, ranging from 1.19 (1.13-1.26) among people aged 20-39, to 0.42 (0.40-0.45) among people aged 60+.

**Table 2.** Odds of any primary care visit by immigration group, stratified by age, British Columbia, Canada, 2019/20-2021/2 (n= 3,707,158)

|  | **Age 20-39** |  | **Age 40-59** |  | **Age 60+** |  |
| --- | --- | --- | --- | --- | --- | --- |
|  | **Unadjusted OR**<br>**(95% CI)** | **Adjusted\***<br>**OR (95% CI)** | **Unadjusted OR**<br>**(95% CI)** | **Adjusted\***<br>**OR (95% CI)** | **Unadjusted OR**<br>**(95% CI)** | **Adjusted\***<br>**OR (95% CI)** |
| **Immigration group (reference is non-immigrant)** |  |  |  |  |  |  |
| Long-term immigrant (5+ years) | 0.89 (0.89, 0.90) | 0.86 (0.85, 0.87) | 0.92 (0.91, 0.93) | 0.92 (0.91, 0.93) | 0.52 (0.51, 0.52) | 0.56 (0.55, 0.57) |
| Recent immigrant (<5 years) | 1.10 (1.08, 1.11) | 1.05 (1.03, 1.06) | 0.81 (0.79, 0.82) | 0.87 (0.84, 0.89) | 0.53 (0.51, 0.56) | 0.58 (0.53, 0.62) |
| Recent immigrant (<5 years, low English) | 1.28 (1.23, 1.33) | 1.19 (1.13, 1.26) | 0.80 (0.77, 0.83) | 0.80 (0.76, 0.85) | 0.38 (0.36, 0.39) | 0.42 (0.40, 0.45) |
| **Year (reference is 2019)** |  |  |  |  |  |  |
| 2020 | 0.77 (0.77, 0.77) | 0.77 (0.76, 0.77) | 0.77 (0.77, 0.78) | 0.78 (0.78, 0.79) | 0.77 (0.77, 0.78) | 0.75 (0.75, 0.76) |
| 2021 | 0.82 (0.81, 0.82) | 0.80 (0.80, 0.81) | 0.87 (0.86, 0.87) | 0.87 (0.86, 0.88) | 0.83 (0.82, 0.83) | 0.79 (0.78, 0.79) |
| **Interaction (immigration group and year, reference is 2019, non immigrant)** |  |  |  |  |  |  |
| Long-term immigrant (5+ years), 2020 | - | 0.91 (0.90, 0.92) | - | 0.89 (0.88, 0.90) | - | 0.95 (0.93, 0.96) |
| Recent immigrant (<5 years), 2020 | - | 0.89 (0.88, 0.91) | - | 0.95 (0.93, 0.98) | - | 0.85 (0.78, 0.92) |
| Recent immigrant (<5 years, low English), 2020 | - | 0.87 (0.82, 0.93) | - | 0.83 (0.78, 0.88) | - | 0.73 (0.68, 0.77) |
| Long-term immigrant (5+ years), 2021 | - | 0.94 (0.93, 0.95) | - | 0.95 (0.94, 0.96) | - | 1.00 (0.98, 1.01) |
| Recent immigrant (<5 years), 2021 | - | 0.91 (0.89, 0.93) | - | 0.95 (0.92, 0.98) | - | 0.96 (0.89, 1.04) |
| Recent immigrant (<5 years, low English), 2021 | - | 0.87 (0.82, 0.93) | - | 0.81 (0.76, 0.87) | - | 0.77 (0.73, 0.82) |

\* Adjusted models also included 5-year age groups, administrative sex, urban/rural residence, neighbourhood income quintile, and Charlson comorbidities

Examining changes over the course of the pandemic, adjusted odds of any primary care visits were lower in 2020/1 compared to 2019/20 (between 0.75 to 0.78 for all age groups, with small variations in the confidence intervals), and rebounded slightly in 2021/2: 0.80 (0.80-0.81) for ages 20-39, 0.87 (0.86-0.88) for ages 40-59, and 0.79 (0.78-0.79) for age 60+, but did not return to pre-pandemic levels. However, interaction terms show that declines in primary care access, during the pandemic, were greater for all immigration groups than for non-immigrants, with the exception of long-term immigrants aged 60+ (1.00, 0.98-1.01); the confidence interval for recent immigrants with high English level aged 60+ intersects with one (0.96, 0.89-1.04) showing no meaningful difference between this group and non-immigrants.

Adjusted odds of access to in-person care (Table 3) showed similar patterns to any primary care visit, with higher adjusted odds for recent immigrants with low English language level among people aged 20-39 (1.28, 1.21-1.35), but lower adjusted odds among people aged 40-59 (0.87, 0.82-0.92) and substantially lower odds among people aged 60+ (0.46, 0.44-0.49). Among people aged 60+ we also saw lower access to in-person care among long-term immigrants (0.58, 0.57-0.59) and recent immigrants with high English level (0.61, 0.57-0.65) compared to non-immigrants.

**Table 3.** Odds of an in-person primary care visit by immigration group, stratified by age, British Columbia, Canada 2019/20-2021/2 (n= 3,707,158)

|  | **Age 20-39** |  | **Age 40-59** |  | **Age 60+** |  |
| --- | --- | --- | --- | --- | --- | --- |
|  | **Unadjusted OR**<br>**(95% CI)** | **Adjusted\***<br>**OR (95% CI)** | **Unadjusted OR**<br>**(95% CI)** | **Adjusted\***<br>**OR (95% CI)** | **Unadjusted OR**<br>**(95% CI)** | **Adjusted\***<br>**OR (95% CI)** |
| **Immigration group (reference is non-immigrant)** |  |  |  |  |  |  |
| Long-term immigrant (5+ years) | 0.92 (0.91, 0.93) | 0.91 (0.90, 0.92) | 0.93 (0.93, 0.94) | 0.97 (0.96, 0.98) | 0.65 (0.64, 0.65) | 0.58 (0.57, 0.59) |
| Recent immigrant (<5 years) | 1.11 (1.10, 1.12) | 1.10 (1.08, 1.12) | 0.82 (0.81, 0.83) | 0.88 (0.86, 0.90) | 0.63 (0.61, 0.66) | 0.61 (0.57, 0.65) |
| Recent immigrant (<5 years, low English level) | 1.33 (1.29, 1.37) | 1.28 (1.21, 1.35) | 0.91 (0.88, 0.94) | 0.87 (0.82, 0.92) | 0.49 (0.48, 0.51) | 0.46 (0.44, 0.49) |
| **Year (reference is 2019)** |  |  |  |  |  |  |
| 2020/1 | 0.30 (0.30, 0.30) | 0.27 (0.27, 0.27) | 0.24 (0.24, 0.25) | 0.22 (0.22, 0.23) | 0.19 (0.19, 0.19) | 0.16 (0.16, 0.16) |
| 2021/2 | 0.37 (0.37, 0.38) | 0.35 (0.35, 0.35) | 0.35 (0.35, 0.36) | 0.34 (0.34, 0.34) | 0.29 (0.29, 0.29) | 0.25 (0.25, 0.26) |
| **Interaction (immigration group and year, reference is 2019, non immigrant))** |  |  |  |  |  |  |
| Long-term immigrant (5+ years), 2020 |  | 0.95 (0.93, 0.96) |  | 0.95 (0.94, 0.96) |  | 1.32 (1.30, 1.34) |
| Recent immigrant (<5 years), 2020 |  | 0.94 (0.92, 0.96) |  | 1.09 (1.06, 1.13) |  | 1.22 (1.13, 1.32) |
| Recent immigrant (<5 years, low English level), 2020 |  | 0.94 (0.89, 1.01) |  | 1.04 (0.98, 1.11) |  | 1.14 (1.07, 1.22) |
| Long-term immigrant (5+ years), 2021 |  | 0.92 (0.90, 0.93) |  | 0.93 (0.92, 0.94) |  | 1.21 (1.19, 1.23) |
| Recent immigrant (<5 years), 2021 |  | 0.89 (0.87, 0.91) |  | 0.95 (0.92, 0.98) |  | 1.08 (1.00, 1.16) |
| Recent immigrant (<5 years, low English level), 2021 |  | 0.91 (0.86, 0.98) |  | 0.92 (0.87, 0.99) |  | 0.99 (0.93, 1.05) |

\* Adjusted models also included 5-year age groups, administrative sex, urban/rural residence, neighbourhood income quintile, and Charlson comorbidities

As expected, the odds of in-person primary care visits were dramatically lower in 2020 compared to 2019 overall, and there was only a slight rebound in 2021/2. Interaction terms (that analyzed the combined effects of immigration groups, stratified by age) showed that among people aged 20-49, declines in adjusted odds of in-person care were greater for all immigration groups compared to non-immigrants, with the exception of recent immigrants with high English level aged 40-59 in 2020 (1.09, 1.06-1.13). Among people ages 60+ declines in odds of in-person care were somewhat more moderate for all immigration groups in 2020 (long-term immigrant (1.32, 1.30-1.34), recent immigrants (1.22, 1.13-1.32), recent immigrants with low English language level (1.14, 1.07-1.22). However, this effect only persisted into 2021/2 among long-term immigrants (1.21, 1.19-1.23).

## Discussion

Our research explored changes in primary care use over the course of the pandemic by age and immigration group. We found that in British Columbia, between 2019/20 and 2021/2, differences in primary care use by immigration group vary by age, with disparities in access particularly apparent among people ages 60+. Within this group, recent immigrants with low English language level had half the odds of any primary care visit compared to non-immigrants. In addition, we observed greater declines in access during the pandemic among immigrants compared to non-immigrants for all immigration groups.

Previous research has documented inequities (ie. differential healthcare access that reflects differences by social position and not by need for healthcare) in access to virtual care among populations with limited digital literacy or access, such as older adults, and those with limited official language proficiency (39). However, our study highlights that this is not compensated for by higher access to in-person care, as might be hoped. While previous research has highlighted inequity in access to healthcare (11,40), our findings troublingly show: persistent and growing inequities by immigration group in the context of COVID-19 among older adults in particular, and growing inequities by immigration group among younger age groups as well.

Our findings suggest that recent immigration and lower English level interact in shaping access to primary care for older adults, a finding also reported in Nouri et al. and Wong et al. (39,41); and, consistent with observations of better access to primary care among long-term immigrants with official language proficiency, and non-immigrants reported in Saskatchewan (42) and nationally (43,44). Both approachability and accessibility of care, and support for care in languages other than English are directly modifiable through responsive policy and service planning. Although previous research (20,45,46) and recent results from Ontario (1) pointed to virtual care as a promising option that could enhance access to primary care during the pandemic, our results showed that, even with the introduction of fee codes that allowed billing health plans for virtual care, access to any primary care, virtual or in-person, has not returned to pre-pandemic levels. Lack of systemic infrastructure supporting newly arrived immigrants and meaningful integration of language support prevented virtual care from being a suitable alternative to in-patient care, at least for immigrants 60+ with low English language level and recent arrival to Canada, consistent with research elsewhere in Canada, Saskatchewan, Alberta (47,48) and the US (49,50). That most people in this group were sponsored family members in our study indicates a particular gap in settlement and health system supports for elder family members reuniting with family in Canada. Interventions to support better access could include: Implementation of interpretation services for all health care providers (to support patients in their language of preference), inclusion of patient navigators (with training in cultural awareness) in healthcare teams, targeted technology support (with physical spaces and in-person navigators), and increased funding for community centers.

A strength of our study is that we used population-based linked administrative data and directly capture features of healthcare and immigration systems that are modifiable, and that structurally determine health (51). Among the potential limitations of our study associated with the use of health administrative data analysis are 1) that the billing codes in British Columbia did not make a clear distinction between video and phone calls for virtual visits, 2) that we examined only non-immigrants and people with permanent immigration status who were continuously registered over the study period, and not temporary or undocumented migrants, who experience more profound barriers to care and are very understudied in Canada (52), and 3) that administrative sex was collected only as a binary set of options (M/F).

## Conclusion

We found evidence of growing inequities by immigration group in access to primary care during the COVID-19 pandemic in BC, particularly for people ages 60 and older. Expanding primary care service delivery that is tailored to meet the needs of recent and long-term im/migrants, and especially older immigrants with low English levels, is needed in order to achieve universal health care access in Canada.

## Data Availability

The data that support the findings of this study are approved for use by data stewards and accessed through a process managed by Population Data BC. The data sets used for this study will be archived, and requests for access to them in the context of verification of study findings can be made to PopData (https://www.popdata.bc.ca/data_access). We are not permitted to share the research extract used in this analysis with other researchers.

## Declarations

### Ethics approval and consent to participate

IRIS holds ethical approval from the Simon Fraser University (SFU) and Providence healthcare/University of British Columbia (UBC) harmonized ethics review boards. Access to data provided by the Data Steward(s) is subject to approval, but can be requested for research projects through the Data Steward(s) or their designated service providers. All inferences, opinions, and conclusions drawn in this publication are those of the author(s), and do not reflect the opinions or policies of the Data Steward(s).

### Consent for publication

Not applicable

### Competing interests

We have no conflicts of interest to declare.

### Funding

This study was funded by the Michael Smith Foundation for Health Research COVID-19 Research Response Fund grant “Preventing and Mitigating the Impacts of COVID-19 Among Im/migrants in British Columbia: Rapid Mixed-Methods Data to Inform Policy and Programmes.” Dr. Wiedmeyer receives salary support through a Trainee Award from Michael Smith Health Research BC.

### Authors’ contributions

Cecilia Sierra-Heredia and Ruth Lavergne conceived of the paper and drafted the manuscript in collaboration with all the co-authors. Shira Goldenberg, Mei-ling Wiedmeyer, and Ruth Lavergne secured funding for this research and provide leadership to broader IRIS study. The analyses were planned in collaboration with the quantitative working group, which includes all co-authors. Sandra Peterson carried out all analyses. All authors contributed to critical interpretation of findings and approved the final manuscript.

## Acknowledgements

We would like to thank IRIS study staff and community partners for their expertise and advice.

## Appendix 1.

**Full regression results**

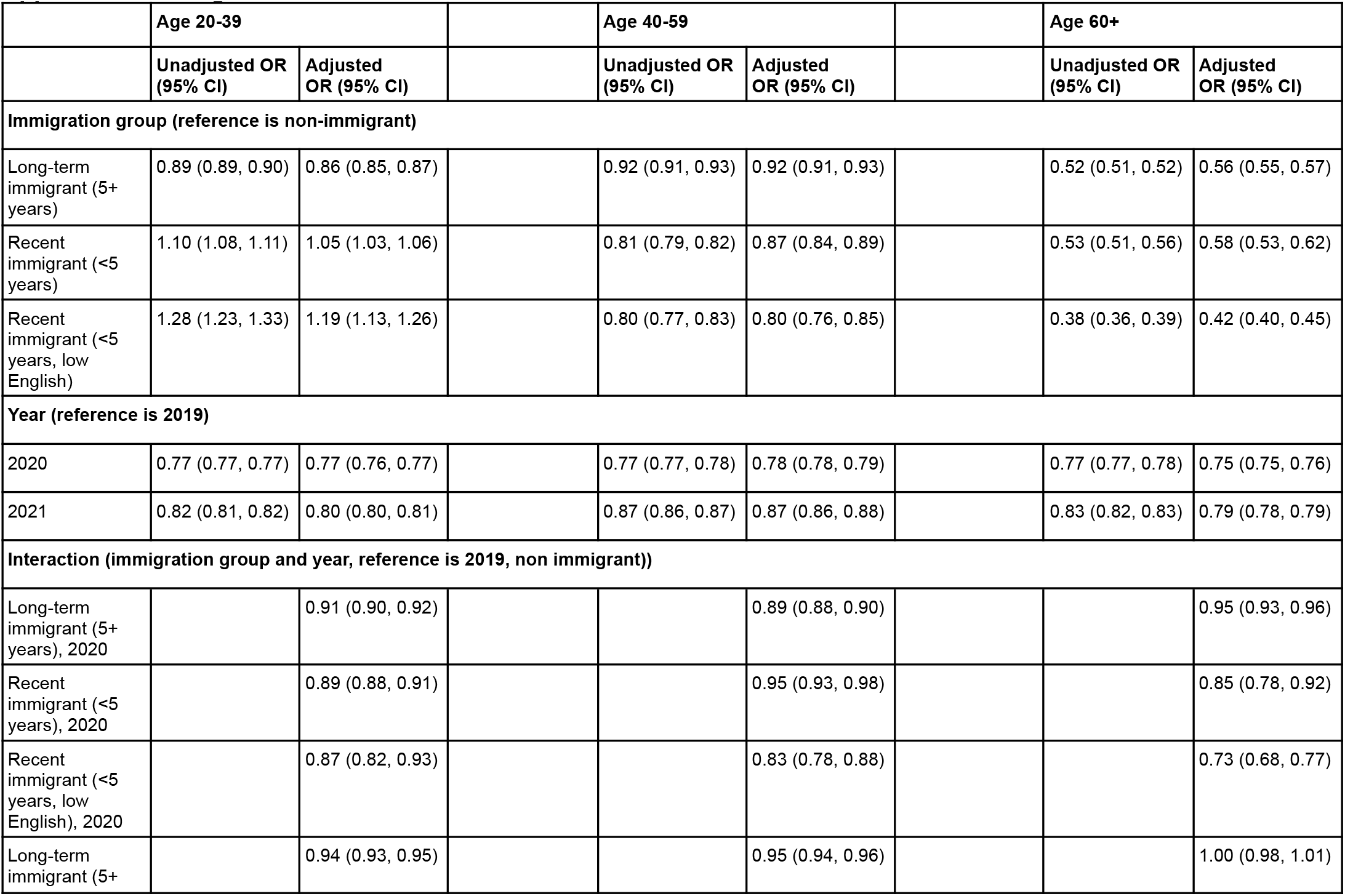
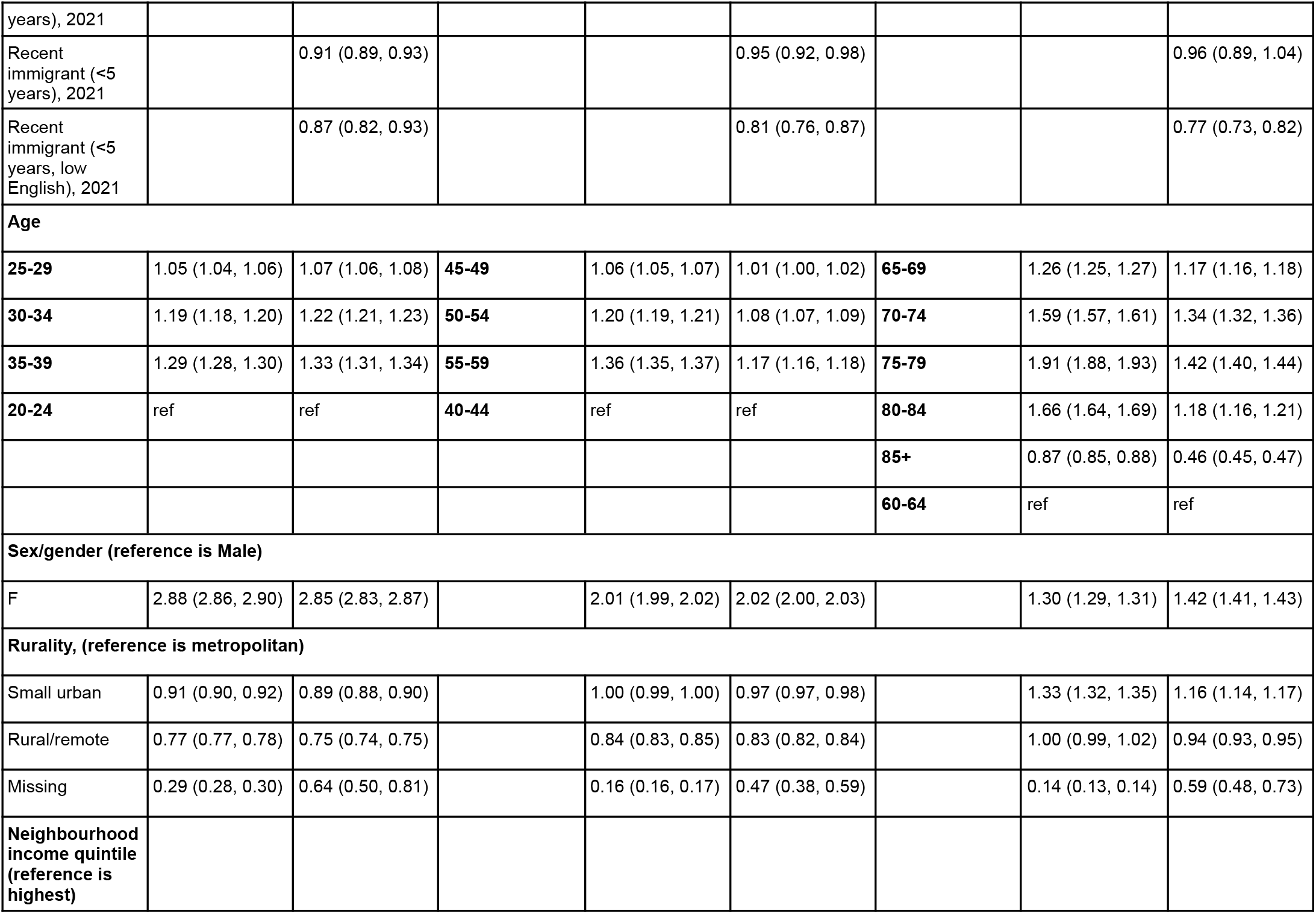
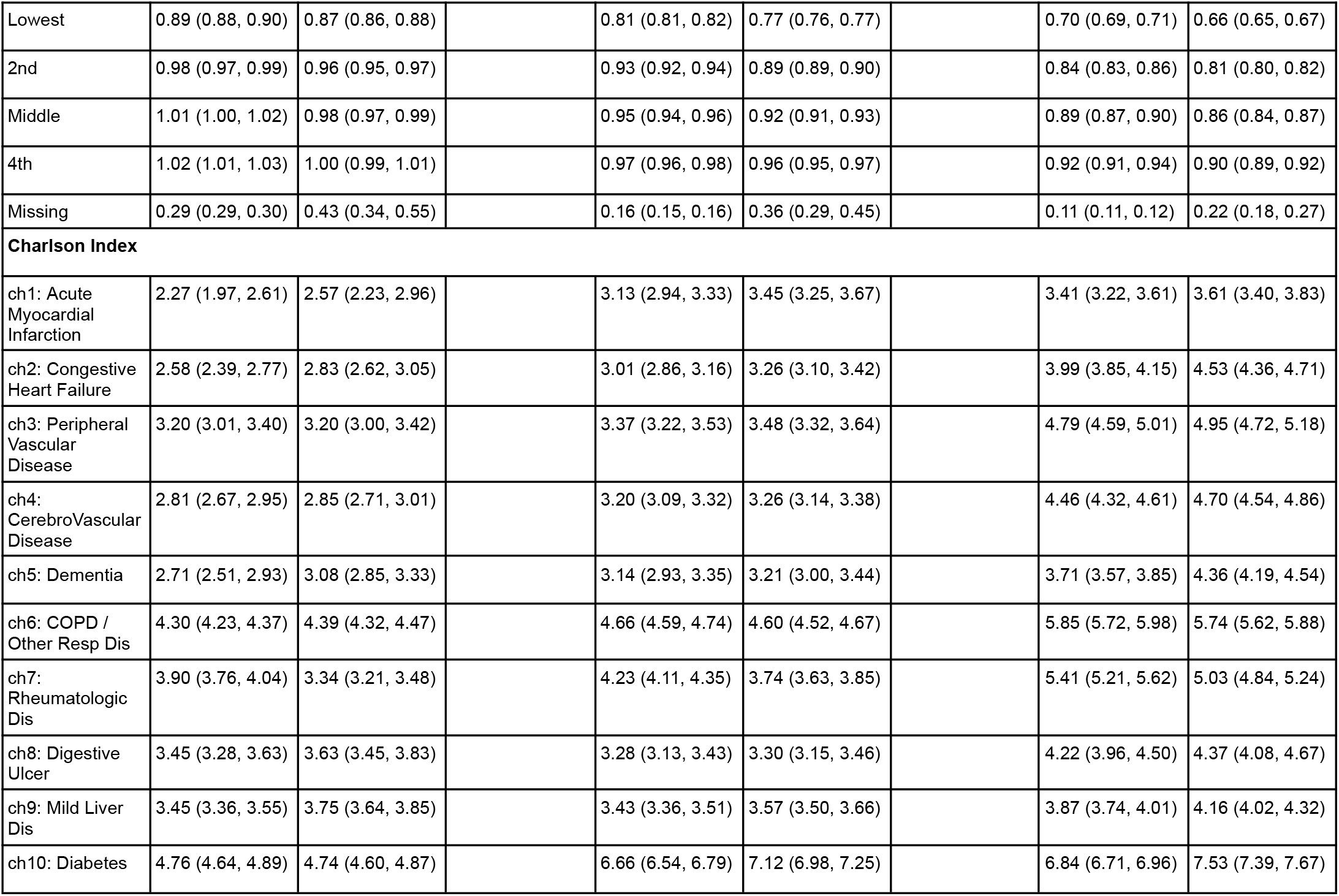
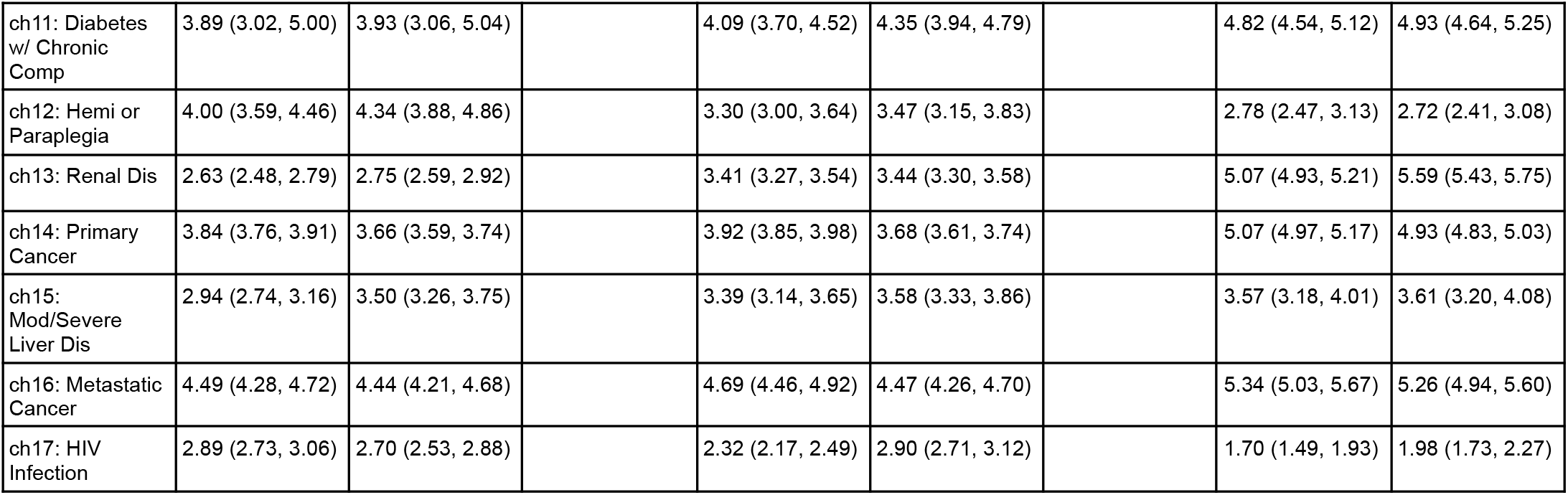

## Notes

### Competing Interest Statement

The authors have declared no competing interest.

### Summary of Updates

Tables and references

